# Electronic health records to test multimorbidity influences to plasma biomarker interpretation for Alzheimer’s disease

**DOI:** 10.1101/2025.08.16.25333799

**Authors:** Katheryn A.Q. Cousins, Rory Boyle, Colleen Morse, Anurag Verma, Christopher A. Brown, Kyra S. O’Brien, Marina Serper, Nadia Dehghani, Penn Medicine BioBank, Corey T. McMillan, Edward B. Lee, Leslie M. Shaw, David A. Wolk

**Author notes:** Correspondence should be addressed to: Katheryn A.Q. Cousins Penn Frontotemporal Degeneration Center Richards Medical Research Laboratories 3700 Hamilton Walk, Philadelphia, PA 19104.

## Abstract

**Objective:** Plasma biomarkers of Alzheimer’s disease (AD) pathology are frequently tested in specialized research settings, limiting generalizability of findings. Using electronic health records and banked plasma, we evaluated plasma biomarkers – phosphorylated tau 217 (p-tau_217_), β-amyloid 1-42/1-40 (Aβ_42_/Aβ_40_) and p-tau_217_/Aβ_42_ – in a real-world, diverse clinical population with multimorbidities.

**Methods:** Participants (n=617; 44% Black/African American; 41% female) were selected from the University of Pennsylvania Medicine BioBank with plasma assayed using Fujirebio Lumipulse. International Classification of Diseases (ICD) Ninth and Tenth Revision codes determined AD dementia (ADD; n=43), mild-cognitive impairment (MCI; n=140), unspecified/non-AD cognitive impairment (CI; n=106), and cognitively normal cases (n=328), and other medical histories. APOE ε4, body mass index (BMI), metrics of kidney function (*e.g.*, eGFR), and liver disease were derived from electronic health records. Multivariable models identified factors related to plasma levels. Previously established cutpoints classified AD status (“AD+”, “AD-”, or “Intermediate”).

**Results:** Plasma p-tau_217_/Aβ_42_had the strongest association with known AD-related factors – MCI, ADD, future progression to MCI/ADD, age, and APOE ε4 – compared to p-tau_217_ and Aβ_42_/Aβ_40_. Plasma p-tau_217_/Aβ_42_ was also associated with eGFR, diabetes, and history of hearing loss. Importantly, AD-related factors were most frequent/severe for AD+ classification by p-tau_217_/Aβ_42_, while medical morbidities were most frequent/severe for Intermediate classification. Exploratory analyses test p-tau_217_/Aβ_42_ adjusted for eGFR to eliminate its influence on plamsa levels.

**Interpretation:** In this real-world dataset, we identified effects of multimorbidities on plasma biomarkers, especially kidney function. The p-tau_217_/Aβ_42_ ratio had low rates of Intermediate classification and may help to account for multimorbidity effects on plasma levels.

## 1. Introduction

Plasma biomarkers have the potential to democratize Alzheimer’s disease (AD) diagnosis, with accumulating evidence of high accuracy to detect AD pathology and strong correlations with β-amyloid (Aβ) and tau pathologies^1–5^. In particular, plasma phosphorylated tau 217 (p-tau_217_) has excellent agreement with amyloid positron emission tomography (PET) and autopsy data^6–8^, and was recently recommended as a core AD biomarker^9^. Combining plasma p-tau_217_ with β-amyloid 1-42 (Aβ_42_) in a ratio (p-tau_217_/Aβ_42_) or in a linear combination with Aβ_42_/Aβ_40_ can further improve AD diagnostic performance, as well as mitigate the influence of non-AD factors^8,10–12^. Indeed, the p-tau_217_/Aβ_42_ratio from Fujirebio Lumipulse recently gained approval from the Food and Drug Administration (FDA)^13^. Still, there are limited studies of plasma AD biomarkers in non-specialty care settings^14–16^. Most testing has been conducted in specialized memory centers, which tend to have higher AD incidence^17,18^, fewer and less severe medical morbidities^19,20^, and be more sociodemographically homogeneous than the general population^15,21^. Thus, testing of blood biomarkers in real-world contexts is needed to interrogate the various factors that can influence blood biomarker performance and interpretation^22–24^.

Here we use electronic health record (EHR) data from the University of Pennsylvania (Penn) Medicine BioBank (PMBB)^25^ to identify factors that can influence AD plasma biomarkers. Plasma concentrations of p-tau_217_, Aβ_42_, and Aβ_40_ were measured using Fujirebio Lumipulse^8,10^. We tested AD-related factors (clinical AD dementia |ADD| or mild cognitive impairment [MCI], APOE ε4 carriers, older age), demographics (*e.g.*, sex, racial identity), and medical history of dementia risk factors, including hearing loss, high LDL cholesterol, depression, traumatic brain injury (TBI), diabetes, hypertension, obesity, and visual loss/impairment^26,27^. We included tests of medical conditions that might affect plasma levels independently of AD pathology, including liver and kidney function and body mass index (BMI)^19,20,28^. To classify individuals for AD pathology (AD+, Intermediate, AD-), we used the recently recommended 2-cutpoint approach^18^ and applied previously established 95% sensitivity and 95% specificity cutpoints^8^. In an exploratory analysis, we calculated plasma levels adjusted for estimated glomerular filtration rate [eGFR] (*i.e.*, kidney function) which can alter blood biomarkers independently of AD pathology^19,20,29^. Finally, we investigated which factors associated with biomarker classifications (*i.e.*, AD+, AD-, or Intermediate).

Real-world EHR data is typically limited in access to gold-standard measures of pathology and formal diagnoses. Nonetheless, we capitalize on the rich breadth of data to identify complex relationships of individual-level factors that influence AD plasma biomarkers. Thus, our objective is not to assess diagnostic accuracy of plasma biomarkers. Instead, we leverage this medically diverse dataset to identify complex relationships of individual-level factors that influence AD plasma biomarkers. Additionally, we take advantage of the converse relationship of higher plasma p-tau_217_ and lower plasma Aβ_42_/Aβ_40_ associated with AD pathology. We hypothesize that known AD-related factors (*e.g.*, APOE ε4, age) will show increased p-tau_217_ and lower Aβ_42_/Aβ_40_, whereas non-AD related plasma factors (*e.g.*, kidney function) may show broad increases or broad decreases across all analytes.

## 2. Methods

Initial selection of 846 plasma samples from the PMBB (https://pmbb.med.upenn.edu/) was based on a random selection of Penn Medicine patients with ICD codes for ADD or MCI at any time and patients without ADD/MCI, matched for age and race. For this study, inclusion criteria were complete data for plasma p-tau_217_ and Aβ_42_ (n=814), age at plasma collection ≥50 years (n=660), Black or White racial identity (n=654), and complete data for metrics of kidney dysfunction (eGFR, creatinine) and BMI, which are known factors associated with plasma biomarker levels^20,30,31^; the result was 617 total samples for this study.

Individual-level data from the PMBB are subject to institutional data use agreements and patient privacy regulations, and access is limited to qualified researchers via the PMBB application process (https://pmbb.med.upenn.edu/investigators.php). Study involving human subjects were approved by the Penn’s Institutional Review Board and informed consent was obtained at study enrollment by study investigators. This cross-sectional observational study follows the Strengthening of Reporting of Observational Studies in Epidemiology (STROBE) guidelines.

### 2.1 Plasma collection and analysis

Blood was collected in EDTA tubes, spun at 1500rcf for 15 minutes at ambient tempurature. The resulting plasma fraction was stored in 1mL aliquots and stored at −80°C. Fujirebio Lumipulse measured plasma concentrations of p-tau_217_, Aβ_42_, and Aβ_40_ according to previous methods^8^.

### 2.2 Cognitive Status

ICD Ninth (ICD-9) and Tenth Revision (ICD-10) Clinical Modification codes are reflected in EHR billing and claims data, and can be used to probe whether a condition is likely present. Nonetheless, we note that ICD codes are not always equivalent to a formal diagnosis, and should be considered evidence of probable presence of a condition^32^.

One or more encounter-level ICD-9 and ICD-10 codes identified likely ADD (331.0, G30.0, G30.1, G30.8, G30.9), MCI (331.83, G31.84), and other/not otherwise specified (NOS) cognitive impairment (CI) (331.11, 331.19, 331.6, 331.82, 780.93, 780.97, 797, 799.[51 – 53, 55, 59], F01.5[11, 18, 2, 3], F01.A[0, 11, 3, 4], F01.B[18, 3], F01.C0, G12.21, G31.[01, 09, 83, 85], R41.[0-4, 81-83, 840-842, 844, 89, 9]). To capture history of a condition, ICD codes were included if they occurred ≤15 years prior to/at plasma collection. In addition, because there is frequently a diagnostic delay associated with MCI/ADD^33,34^, we included cognitive ICD codes up to one year after plasma collection. If CI was absent (0 ICD codes for ADD, MCI and Other/NOS CI), patients were considered cognitively unimpaired/normal.

#### Future progression to MCI/ADD

ICD codes ≥1 year(s) after plasma collection stratified persons who progressed to MCI or ADD in the future (≥1 MCI/ADD code) *vs.* stable/no progression (0 codes). Normal and Other/NOS CI could progress to either MCI or ADD, and persons with MCI could progress to ADD; ADD persons could not progress.

### 2.3 Medical morbidities

#### 2.3.1 Histories of morbidities

Age, sex, race, and histories of smoking and alcohol use were collected from the EHR and social history; passive/second-hand smoke was considered as having a history of smoking. In addition, APOE ε4 status was determined based on rs429358-C allele counts from Whole Exome Sequencing data.

ICD-9 and ICD-10 codes identified likely histories of depression, diabetes, hypertension, stroke/cerebrovascular disease^35^, and TBI^36^. ICD-9 and ICD-10 codes were determined by PheCodes (https://phewascatalog.org/phewas/#phex) for visual loss (PheCode=Blindness and low vision, Constriction of visual field, Disorders of optic nerve and visual pathways, Disorders of visual cortex, Visual distortions and subjective visual disturbances, or Visual field defects; Category=Sense Organs), and hearing loss (PheCode=Hearing Impairment; Category=Sense Organs). ICD codes were included if they occurred ≤15 years prior to/at plasma collection, as well as ≤1 years after plasma collection to allow for a diagnostic delay. To increase specificity, a person was considered likely positive for a medical morbidity if 2 or more ICD encounters were associated with that condition.

#### 2.3.2 Blood tests/BMI

Blood tests were taken closest to plasma sample with a mean interval of 0.06 years (standard deviation [SD]=0.41; minimum= −4.98; maximum=1).

##### Kidney

Chronic kidney disease (CKD) has been previously associated with individual plasma AD analytes (but not the Aβ_42_/Aβ_40_ ratio), independent of AD pathology^19,20^. We therefore tested three measures of kidney function to examine which associated with AD plasma biomarkers: creatinine (mg/dL), blood urea nitrogen [BUN] (mg/dL), and eGFR (mL/min/1.73 m^2^)^37^. Creatinine and BUN are waste products filtered out of the blood when kidney function is healthy, and eGFR is calculated based on serum creatinine, age, and sex. For creatinine, one outlier of 328.9 was excluded. Our sample includes a broad range of kidney function measured by creatinine (range=0.36 – 28), BUN (range=2 – 116), and eGFR (range = 0.062 – 119.766). 187 patients had moderate/Stage 3 CKD or worse (eGFR<60) and 48 exhibited severe/Stage 4+ CKD (eGFR<30).

##### Liver

Alanine aminotransferase (ALT) – an enzyme found in the liver – and total bilirubin – a breakdown product of red blood cells – tested associations of liver function with AD plasma biomarkers^28^. Our sample includes a broad range of liver function/inflammation (ALT range=3 – 230; Bilirubin range=0.2 – 12.4).

##### Obesity

Body mass index (BMI) was computed as a measure of obesity, which may also affect AD plasma biomarker levels^20^ as well as being a risk factor for dementia^26^. Our sample includes a broad range of BMI (BMI range=15 – 57). 223 patients were overweight (BMI≥30).

### 2.4 Statistical Analyses

Variables were not normally distributed; non-parametric tests (*e.g.*, Mann-Whitney-Wilcoxon, Kruskal-Wallis, Spearman’s rho) were used in unadjusted comparisons, reported in Figures. For clearer visualization, extreme values were Winsorized to 95 or 99^th^ percentile where noted in Figures.

In linear and logistic regression models and Pearson correlations, plasma AD biomarkers, creatinine, BMI, LDL cholesterol, and BUN were log-transformed, and all continuous variables were scaled to standardize β-coefficients and allow for comparisons of effect size across variables; β-estimates or odds ratios (OR), 95% confidence intervals (95%CI), and *p*-values were reported. Significance threshold was α=0.05. Analyses were conducted using R version 4.4.0 (2024-04-24) software. R code can be made available upon reasonable request to authors.

#### Multicollinearity

As expected, metrics of kidney function were highly correlated: creatinine was highly correlated with eGFR (rho= −0.9, *p*=3.7e-223) and BUN (rho=0.62, *p*=3.1e-66); likewise, BUN and eGFR were correlated with each other (rho= −0.61, *p*=3.8e-64). Liver metrics ALT and bilirubin were also correlated (rho=0.19, *p*=0.0000077). Variance inflation factor (VIF) therefore assessed issues of multicollinearity in models. For all plasma biomarkers (p-tau_217_/Aβ_42_, p-tau_217_, Aβ_42_/Aβ_40_), collinearity was confirmed between creatinine (VIF=9.5) and eGFR (VIF=9.3). All other factors had a VIF<4 (BUN VIF=2.3; all other VIF≤1.9). Therefore creatinine was not included in models.

#### Missing data

To maintain sample size, factors with >100 missing observations (*e.g.*, LDL cholesterol) were not included in multivariable models. When these data were included in models, they were not significantly associated with plasma AD biomarker levels (data not shown). However, we did evaluate their effect on MCI/ADD incidence and biomarker classifications. In addition, 15 cases were missing Aβ_40_ data. Missing data are reported in Table 1.

**Table 1:** Demographic and clinical characteristics of Penn Medicine BioBank (PMBB) participants. For continuous variables, median and interquartile range (IQR) are reported; Kruskal-Wallis tests performed group comparisons. For categorical variables, count (percentage [%]) are provided; chi-square tests performed frequency comparisons. *p*-values are reported for group comparisons.

| Normal | Other/NOS CI | MCI | ADD | p | Missing |
| --- | --- | --- | --- | --- | --- |

**Table 1: Demographic and clinical characteristics of Penn Medicine BioBank (PMBB) participants.**
|  | Normal | Other/NOS CI | MCI | ADD | p | Missing |
| --- | --- | --- | --- | --- | --- | --- |
| n | 328 | 106 | 140 | 43 |  |  |
| Sex = Female (%) | 128 (39.0%) | 47 (44.3%) | 57 (40.7%) | 19 (44.2%) | 0.758 | -- |
| Race = Black (%) | 141 (43.0%) | 53 (50.0%) | 60 (42.9%) | 15 (34.9%) | 0.365 | -- |
| Ethnicity = Hispanic or Latino (%) | 3 (0.9%) | 0 | 0 | 0 | 0.449 | 3 |
| Education | 17.5 [12.8, 18.0] | 14.0 [12.0, 16.0] | 16.0 [12.0, 18.0] | 18.0 [12.0, 18.0] | 0.036 | 467 |
| Age at plasma (years) | 66.2 [59.2, 73.6] | 70.5 [64.1, 75.3] | 68.2 [60.2, 77.5] | 78.0 [72.5, 80.9] | <0.001 | -- |
| APOE $\epsilon 4 = \geq 1$ alleles (%) | 102 (31.1%) | 33 (31.1%) | 54 (38.6%) | 21 (48.8%) | 0.068 | -- |
| ALS (%) | 0 | 0 | 1 (0.7%) | 0 | 0.332 | -- |
| BMI | 28.0 [24.0, 32.0] | 27.0 [24.0, 31.0] | 27.0 [24.0, 31.2] | 26.0 [22.0, 29.5] | 0.123 | -- |
| eGFR | 75.5 [57.8, 90.4] | 69.1 [49.1, 85.7] | 72.8 [56.4, 90.3] | 72.0 [55.3, 79.7] | 0.162 | -- |
| Creatinine | 1.0 [0.8, 1.3] | 1.0 [0.8, 1.4] | 1.0 [0.8, 1.3] | 1.0 [0.9, 1.2] | 0.677 | 1 |
| BUN | 17.0 [13.0, 22.0] | 19.0 [14.0, 25.8] | 18.0 [15.0, 24.2] | 17.0 [14.0, 22.0] | 0.140 | 1 |
| Bilirubin | 0.6 [0.4, 0.8] | 0.5 [0.4, 0.7] | 0.6 [0.4, 0.8] | 0.6 [0.4, 0.8] | 0.555 | 42 |
| ALT | 19.0 [14.0, 27.0] | 16.0 [11.0, 24.0] | 17.0 [12.0, 24.0] | 16.0 [12.2, 22.5] | 0.123 | 39 |
| LDL Cholesterol | 89.0 [68.0, 116.0] | 87.0 [71.2, 114.8] | 83.5 [63.0, 112.2] | 95.0 [73.0, 114.0] | 0.419 | 189 |
| Depression (%) | 47 (14.3%) | 34 (32.1%) | 54 (38.6%) | 3 (7.0%) | <0.001 | -- |
| TBI (%) | 17 (5.2%) | 20 (18.9%) | 21 (15.0%) | 3 (7.0%) | <0.001 | -- |
| Hypertension (%) | 247 (75.3%) | 88 (83.0%) | 126 (90.0%) | 36 (83.7%) | 0.002 | -- |
| Stroke/cerebrovascular (%) | 64 (19.5%) | 30 (28.3%) | 61 (43.6%) | 15 (34.9%) | <0.001 | -- |
| Alcohol (%) | 168 (53.0%) | 48 (45.7%) | 55 (39.9%) | 19 (46.3%) | 0.069 | 16 |
| Smoking (%) | 188 (58.4%) | 75 (70.8%) | 80 (57.1%) | 26 (61.9%) | 0.113 | 7 |
| Diabetes (%) | 106 (32.3%) | 44 (41.5%) | 58 (41.4%) | 15 (34.9%) | 0.163 | -- |
| VisionLoss (%) | 18 (5.5%) | 12 (11.3%) | 19 (13.6%) | 3 (7.0%) | 0.021 | -- |
| HearingLoss (%) | 67 (20.4%) | 34 (32.1%) | 47 (33.6%) | 8 (18.6%) | 0.005 | -- |

#### 2.4.1 Medical and demographic associations with ADD/MCI

Logistic regression tested each modifiable medical and demographic factors associated with MCI or ADD ICD codes, compared to normal cognition or Other/NOS CI. Nominal and Bonferroni corrected *p*-values are reported.

#### 2.4.2 AD plasma biomarker levels

Second, linear models tested factors associated with plasma AD biomarkers. To understand multifactorial contributions to plasma biomarkers and AD pathology, we report both univariable and multivariable relationships with each biomarker.

##### Calculating eGFR-adjusted plasma AD biomarker levels

Models identified eGFR as a large contributing factor in higher concentrations for all plasma analytes (p-tau_217_, Aβ_42_, Aβ_40_); eGFR had an effect size second only to ADD for p-tau_217_, and had the largest effect size for Aβ_42_ and Aβ_40_. In a posthoc, exploratory analysis, we calculated plasma levels adjusted for eGFR. From the PMBB, we identified an independent set of younger adults (n=131; 71 female [54%]; 80 Black and 1 multiracial [62%]), who were aged ≤50 years (mean=38.7, standard deviation [SD]=8.2). Models identified relationships between eGFR and log-transformed p-tau_217_ and p-tau_217_/Aβ_42_(Equations 1 & 2). Trained models were applied to the current data set to calculate eGFR-adjusted plasma levels. While eGFR was associated with both Aβ_42_ and Aβ_40_, it did not influence Aβ_42_/Aβ_40_ levels in multivariable models (seen previously^20^) and so the Aβ_42_/Aβ_40_ ratio was unadjusted. Linear models for plasma p-tau_217_/Aβ_42_ and p-tau_217_ were then repeated using eGFR-adjusted plasma values.

1. *log(ptau_217_) = −0.63 + −0.02 eGFR*
2. *log(ptau_217_/A*β*_42_) = −4.55 + −0.01 eGFR*

We note that eGFR-adjusted plasma values were used across the full range of eGFR, including those with high/normal eGFR values. Thus, eGFR-adjusted plasma values were decreased for those with high eGFR.

To test an alternative approach, a multivariable model tested limiting adjustment to only individuals with low/abnormal eGFR, however eGFR remained significant for p-tau_217_ (β= −0.14, 95%CI= −0.24 – −0.04, p=0.007). By contrast, our initial approach of adjusting by eGFR for all individuals eliminated significant effects of creatinine, eGFR, and BUN on plasma p-tau_217_ and p-tau_217_/Aβ_42_(see results below); this was our final model solution.

#### 2.4.3 Biomarker Classifications

Finally, previously established 95% sensitivity and 95% specificity cutpoints^8^ were applied and classified individuals as AD+, AD-, or Intermediate. Because cutpoints were not derived using eGFR-adjusted values, plasma values for classification used unadjusted values. To compare proportion of Intermediates for each biomarker, we used the test of equal proportions. Chi-square and Kruskal-Wallis tests tested which medical and demographic factors were associated with plasma AD biomarker classifications. We expected known AD associated factors – MCI/ADD, progression to MCI/ADD, age, and APOE ε4 genotype – to be associated with AD+ classification.

## 3. Results

Demographics and characteristics of the data set, stratified by cognitive status, are summarized in Table 1. One person with MCI also had ICD codes indicating amyotrophic lateral sclerosis (ALS; 335.20, G12.21), which can elevate plasma p-tau levels^38^; this person had p-tau_217_/Aβ_42_ of 0.004 (AD-status^8^), p-tau_217_ of 0.145 (Intermediate status^8^), and Aβ_42_/Aβ_40_ of 0.0943 (Intermediate status^8^).

### 3.1 Morbidity associations with CI status

Univariable models examined morbidity factors associated with MCI or ADD diagnosis. MCI/ADD was more frequent in people with histories of depression (OR=2.00, 95%CI=1.30 – 2.90, *p*=0.00076; Bonferroni-*p*=0.011), stroke/cerebrovascular disease (OR=2.60, 95%CI=1.80 – 3.70, *p*=0.00000068; Bonferroni-*p*=0.00001), and hypertension (OR=2.30, 95%CI=1.40 – 3.90, *p*=0.0014; Bonferroni-*p*=0.022). Several associations did not survive correction for multiple comparisons, including APOE ε4 (OR=1.50, 95%CI=1.10 – 2.20, *p*=0.018; Bonferroni-*p*=0.28), alcohol use (OR=0.67, 95%CI=0.47 – 0.96, *p*=0.028; Bonferroni-*p*=0.41) and visual loss (OR=1.80, 95%CI=1.00 – 3.30, *p*=0.039; Bonferroni-*p*=0.59). Race, sex, ALT, creatinine, eGFR, BUN, LDL cholesterol, BMI, and history of diabetes, TBI and hearing loss were not associated with MCI/ADD (BMI: OR=0.85, 95%CI=0.71 – 1.00, *p*=0.065; hearing loss: OR=1.40, 95%CI=0.96 – 2.10, *p*=0.077; TBI: OR=1.60, 95%CI=0.93 – 2.80, *p*=0.083; all other *p*≥0.17).

### 3.2 eGFR and plasma biomarkers

We tested several indicators of kidney function – eGFR, creatinine, and BUN – which have been shown to correlate with plasma biomarkers independently of AD pathology^19,20^. Analyses focused on eGFR as an indicator of kidney function.

Correlations were strong between eGFR and plasma biomarkers in both an independent, reference data set of young adults (Figure 1A) and our PMBB data set (Figure 1B). Indeed, in young adults with low eGFR (>60 mL/min/1.73 m^2^; normal/mild CKD), eGFR was still correlated with log-transformed plasma p-tau_217_ (r= −0.33, 95%CI= −0.48 – −0.16, p=0.00031) and p-tau_217_/Aβ_42_ (r= −0.27, 95%CI= −0.43 – −0.09, p=0.0041). In an exploratory analysis, we computed plasma p-tau_217_ and p-tau_217_/Aβ_42_ levels adjusted for eGFR (Figure 1C).

**Figure 1.**
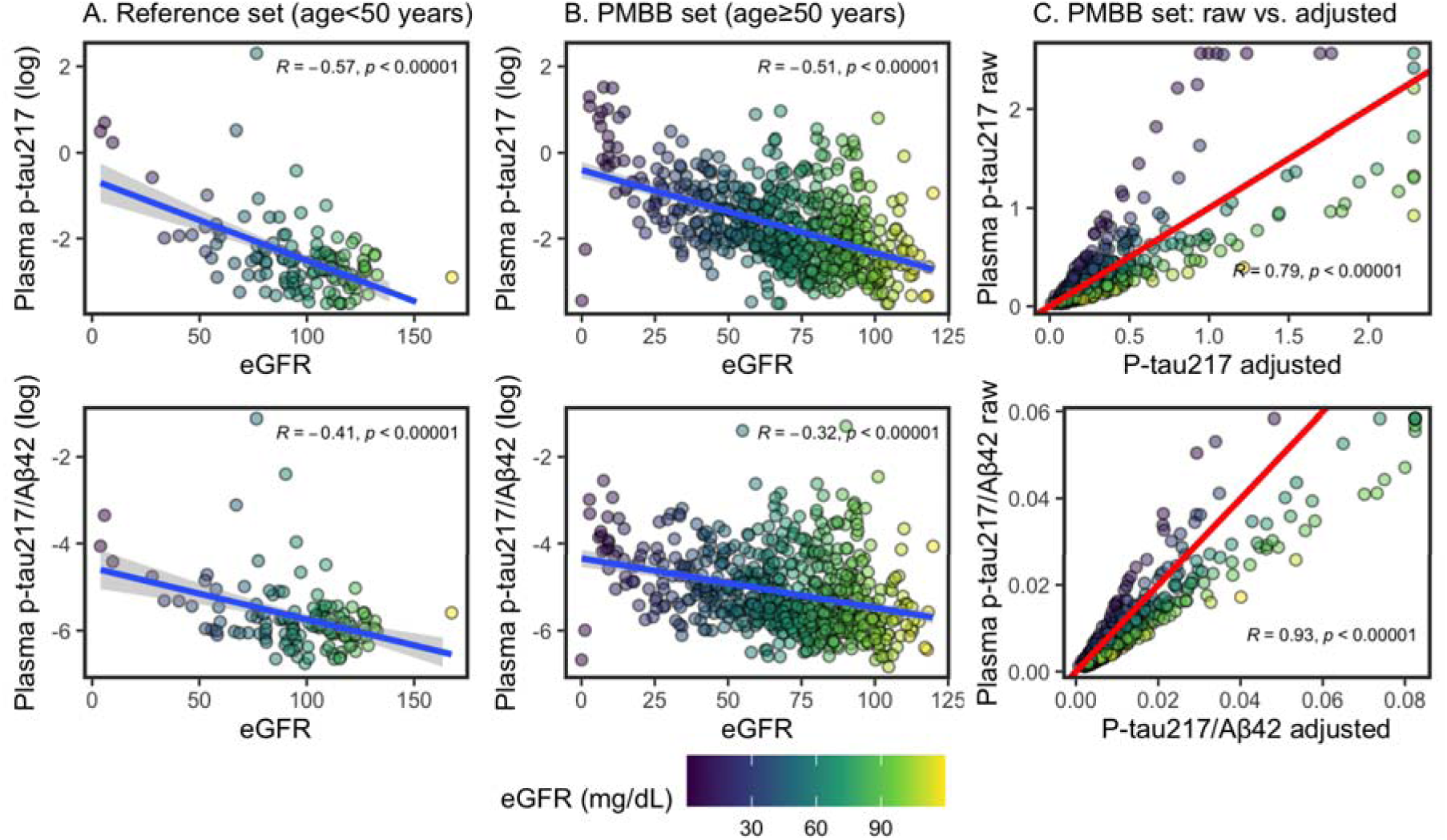
Correlations between plasma biomarkers and creatinine. Color of points indicates eGFR (mL/min/1.73 m^2^). Pearson’s correlations are reported. (**A.**) Relationship between eGFR and log-transformed plasma p-tau_217_ and p-tau_217_/Aβ_42_ in reference dataset of younger adults aged less than 50. Least squares regression line is in blue. Models were developed based on these for p-tau_217_ [*log(ptau_217_) = −0.63 + −0.02 eGFR*] and p-tau_217_/Aβ_42_ [*log(ptau_217_/A*β*_42_) = −4.55 + −0.01 eGFR*]. (**B.**) Relationship between eGFR and log-transformed plasma p-tau_217_ and p-tau_217_/Aβ_42_ in PMBB dataset of older adults. Least squares regression line is in blue. (**C.**) In PMBB dataset of older adults, relationship between raw values and eGFR-adjusted values, based on models derived in young-adults. High values are Winsorized to 99% of mean. Red, diagonal line indicates correlation of 1 (slope=1, intercept=0): adjusted values above the line are lower than raw, and values below are higher than raw.

### 3.3 Plasma biomarker correlates and classifications

Univariable and multivariable linear models tested the relationship between cognition, future progression to MCI/ADD, demographics (race, sex, age), APOE ε4, and medical morbidities.

#### 3.3.1 Plasma p-tau_217_/Aβ_42_

##### Concentrations

Figure 2A shows plasma p-tau_217_/Aβ_42_ concentrations by cognitive diagnosis and future progression to MCI or ADD. In multivariable models (Figure 3B), higher p-tau_217_/Aβ_42_was associated with AD-related factors – diagnosis of MCI (β=0.23, 95%CI=0.02 – 0.44, p=0.033) or ADD (β=0.79, 95%CI=0.46 – 1.12, p<0.001), future progression to MCI/ADD (β=0.33, 95%CI=0.15 – 0.51, p<0.001), older age (β=0.26, 95%CI=0.18 – 0.34, p<0.001), and APOE ε4 carriers (β=0.42, 95%CI=0.27 – 0.57, p<0.001) – as well as lower eGFR (β= −0.23, 95%CI= −0.34 – −0.12, p<0.001) and a history of diabetes (β=0.17, 95%CI=0.00 – 0.33, p=0.049). While not significant in univariable models, hearing loss was significant in multivariable models (β= −0.18, 95%CI= −0.35 – −0.01, p=0.034). BUN (β=0.26, 95%CI=0.18 – 0.34, p<0.001) was significant in univariable models only (Figure 3A). Multivariable model results were consistent for eGFR-adjusted plasma p-tau_217_/Aβ_42_, except that eGFR and diabetes were no longer significant (Figure 3D).

**Figure 2.**
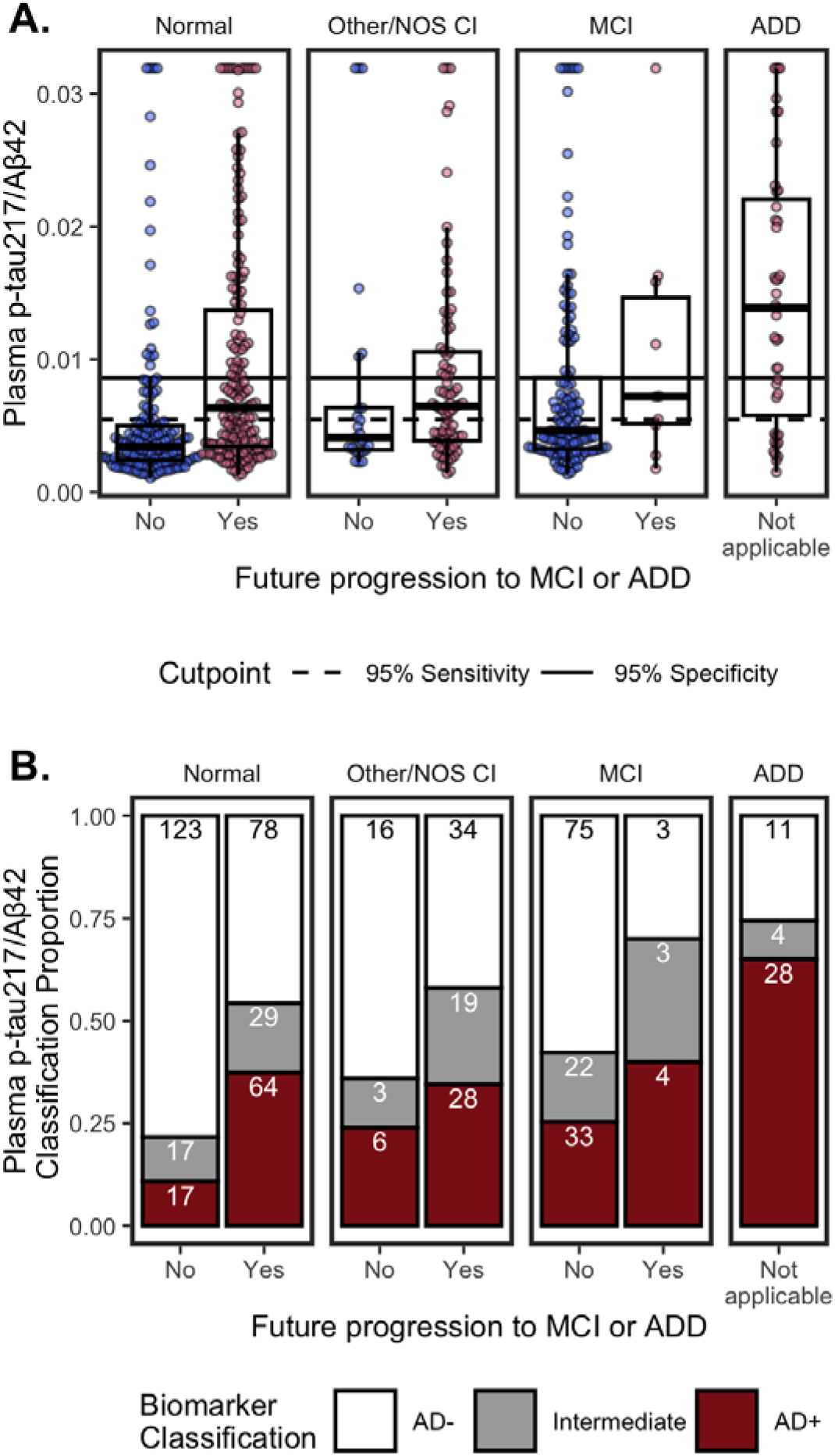
Plasma p-tau_217_/Aβ_42_ by cognitive status (Normal, Other/NOS CI, MCI, ADD) and future clinical progression. (**A.**) Boxplots show median, interquartile range (IQR), and outliers for each plasma biomarker; high plasma levels are Winsorized to 95^th^ percentile for visualization. Color indicates future progression to MCI/ADD (red) or not (blue). Panels show cognitively Normal, Other/NOS CI, MCI, and ADD. Solid horizontal line indicates 0.95 specificity threshold; broken horizonal lines indicate 0.95 sensitivity threshold. (**B.**) Classification of p-tau_217_/Aβ_42_ thresholds by cognitive status (Normal, Other/NOS CI, MCI, ADD) and future clinical progression. Barplots show proportion of classifications as Amyloid-(white), Intermediate (grey), or Amyloid+ (red). Count for each classification are labeled in bars.

**Figure 3.**
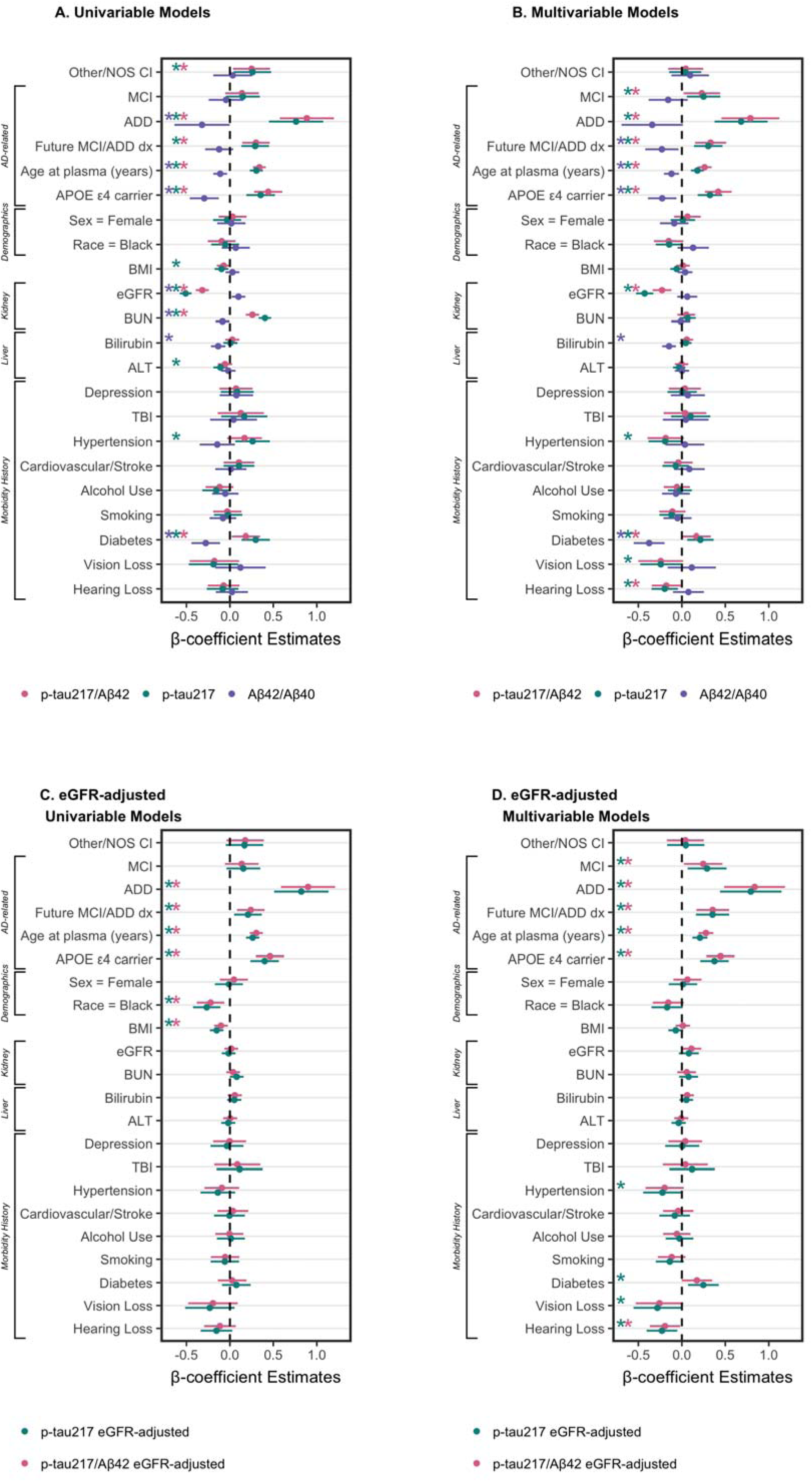
Plasma biomarkers by demographics and morbidities. Univariable (**A., C.**) and multivariable (**B., D.**) linear models of p-tau_217_/Aβ_42_(pink), p-tau_217_ (green), and Aβ_42_/Aβ_40_ (purple), log-transformed and scaled. Models test unadjusted plasma values (**A., B.**) and eGFR-adjusted values (**C., D.**). Dot-and-whisker plots β coefficients and 95% confidence intervals for each predictor (scaled, and log-transformed where appropriate). Asterisks by variable name indicate *p*<0.05.

##### Classifications

Using previously established cutpoints^8^, p-tau_217_/Aβ_42_ classifications were 180 (29%) AD+, 340 (55%) AD-, and 97 (16%) Intermediate (Figure 2B). Because established cutpoints are not eGFR-adjusted, unadjusted p-tau_217_/Aβ_42_ values were used. In line with having AD pathology, AD+ classifications were more likely to have MCI or ADD, progression to MCI/ADD in the future, be older, and have ≥1 APOE ε4 alleles, compared to AD- and Intermediates (Table 2).

**Table 2.**
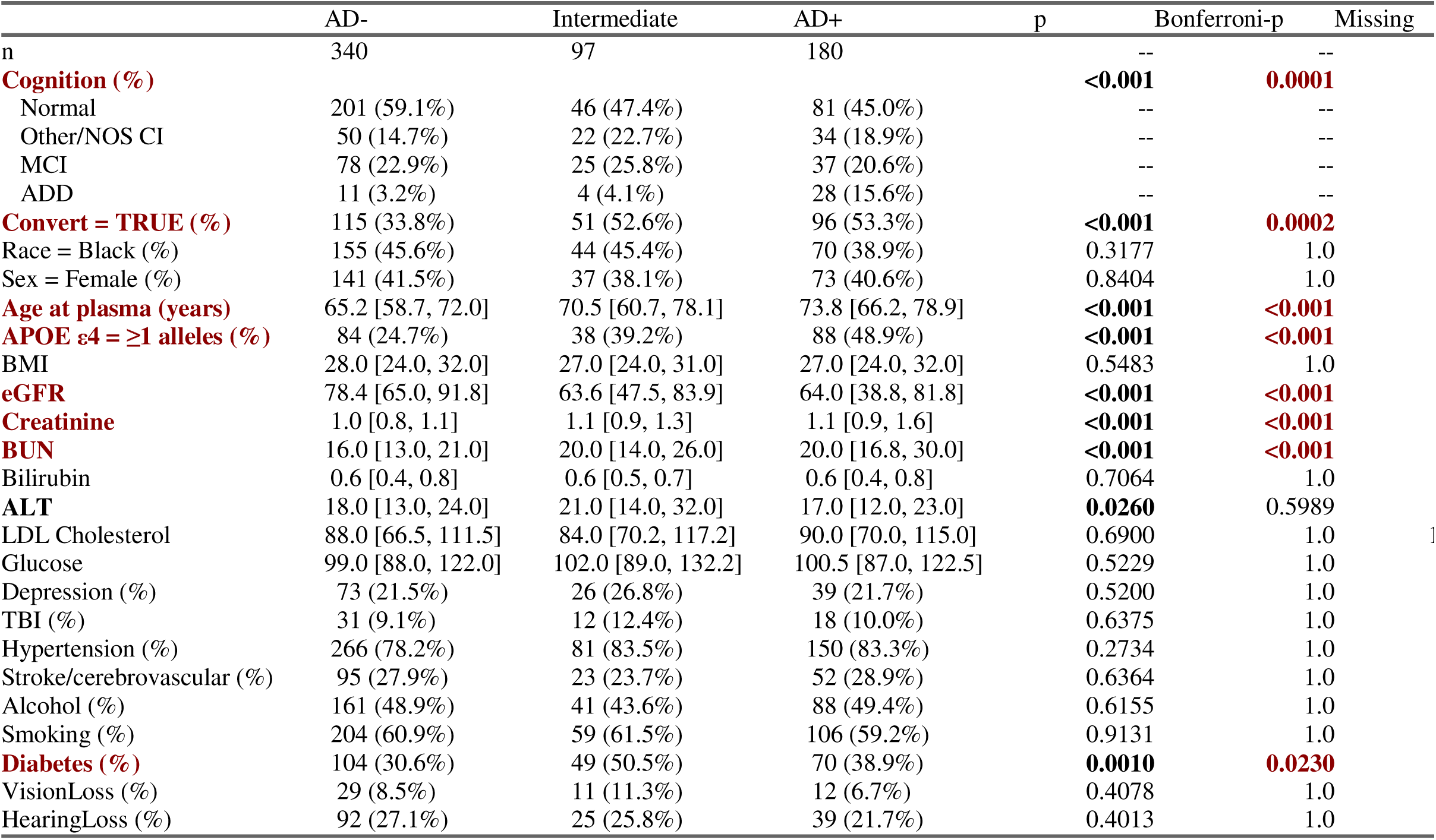
Demographic and morbidity factors that affect plasma p-tau_217_. /Aβ_42_ **classifications**. Summary statistics across AD negative, Intermediate, and positive classifications. Kruskal-Wallis and Chi-square tested group differences, *p*-values and Bonferroni-*p* are reported: *p*<0.05 are bolded and Bonferroni-*p*<0.05 are in red.

Of note, Intermediate status for p-tau_217_/Aβ_42_ was high for co/multimorbidities compared to AD-status (Table 2). Intermediates had the lowest median eGFR, highest ALT and rates of diabetes compared to AD- and AD+. Intermediates and AD+ had lower median creatinine and BUN than AD-.

#### 3.3.2 Plasma p-tau_217_

##### Concentrations

Figure 4A shows plasma p-tau_217_ concentrations by cognitive diagnosis and future progression to MCI or ADD. In multivariable models (Figure 3B), higher p-tau_217_ was associated with MCI (β=0.25, 95%CI=0.06 – 0.44, p=0.011), ADD (β=0.68, 95%CI=0.38 – 0.99, p<0.001), future progression to MCI/ADD (β=0.30, 95%CI=0.14 – 0.47, p<0.001), older age (β=0.18, 95%CI=0.11 – 0.25, p<0.001), APOE ε4 (β=0.32, 95%CI=0.18 – 0.46, p<0.001), lower eGFR (β= −0.43, 95%CI= −0.53 – −0.33, p<0.001), and histories of diabetes (β=0.21, 95%CI=0.06 – 0.37, p=0.006) and hypertension (β= −0.19, 95%CI= −0.38 – −0.00, p=0.048). Histories of hearing loss (β= −0.20, 95%CI= −0.35 – −0.05, p=0.011) and visual loss (β= −0.24, 95%CI= −0.48 – −0.01, p=0.045) were associated with lower p-tau_217_ in the multivariable model, but not univariable models. Conversely, univariable relationships (Figure 3A) with BMI (β= −0.10, 95%CI= −0.17 – −0.02, p=0.017), BUN (β=0.40, 95%CI=0.33 – 0.48, p<0.001), and ALT (β= −0.11, 95%CI= −0.19 – −0.03, p=0.008) were not significant in multivariable models. Multivariable model results were consistent for eGFR-adjusted plasma p-tau_217_, except that eGFR was no longer a significant factor (Figure 3D).

**Figure 4.**
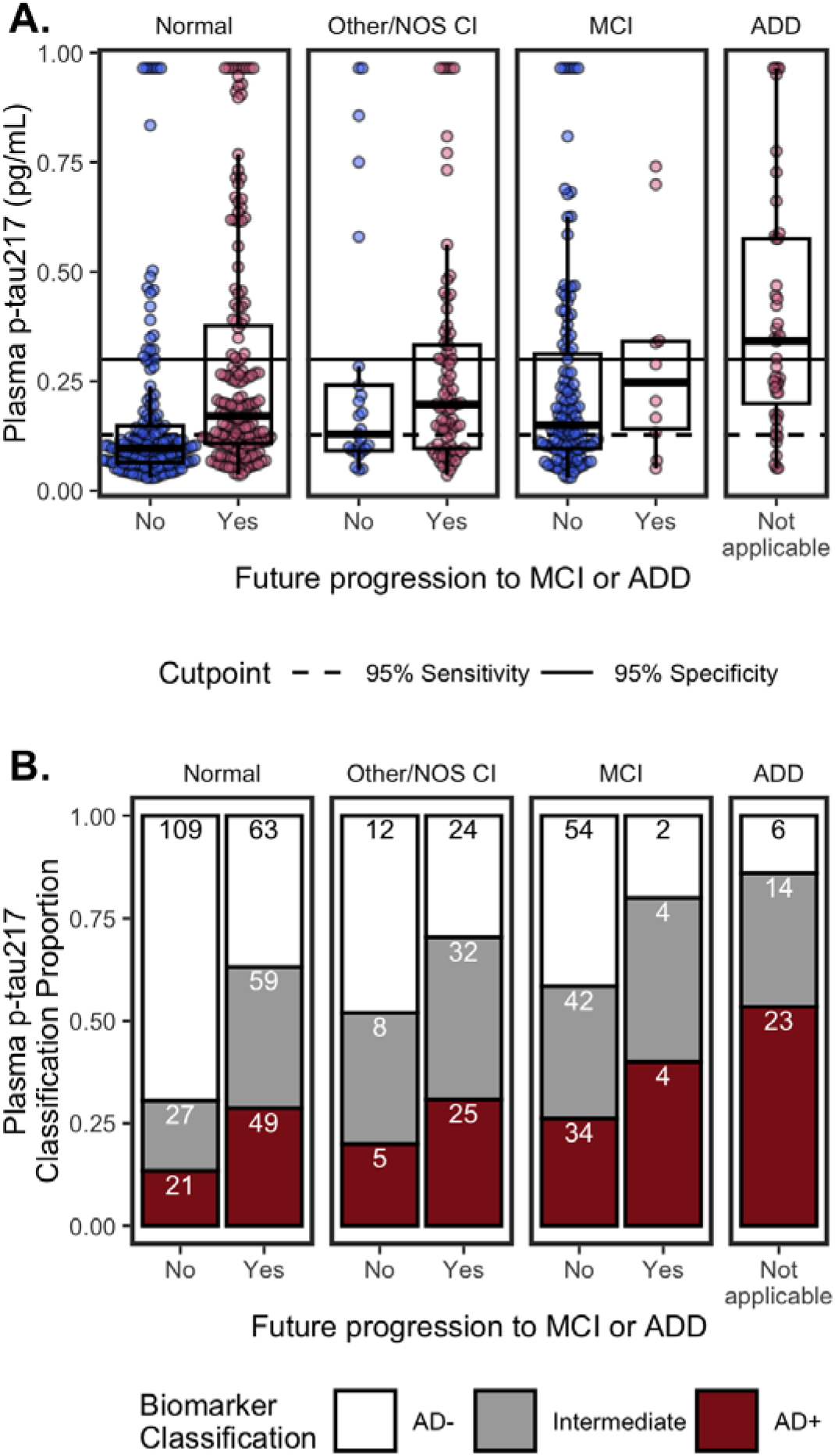
Plasma p-tau_217_ by cognitive status (Normal, Other/NOS CI, MCI, ADD) and future clinical progression. (**A.**) Boxplots show median, interquartile range (IQR), and outliers for each plasma biomarker; high plasma levels are Winsorized to 95^th^ percentile for visualization. Color indicates future progression to MCI/ADD (red) or not (blue). Panels show cognitively Normal, Other/NOS CI, MCI, and ADD. Solid horizontal line indicates 0.95 specificity threshold; broken horizonal lines indicate 0.95 sensitivity threshold. (**B.**) Classifiation of p-tau_217_ thresholds by cognitive status (Normal, Other/NOS CI, MCI, ADD) and future clinical progression. Barplots show proportion of classifications as Amyloid-(white), Intermediate (grey), or Amyloid+ (red). Count for each classification are labeled in bars.

##### Classifications

Using previously established cutpoints^8^, p-tau_217_ classifications were 161 (26%) AD+, 270 (44%) AD-, and 186 (30%) Intermediate (Figure 4B). The percentage of Intermediate cases was significantly higher for p-tau_217_ (χ2(1)=35.51, 95%CI=0.1 – 0.19, p=2.5e-09) than p-tau_217_/Aβ_42_ (16%).

Consistent with having AD pathology, AD+ classifications were more likely to have MCI or ADD, progress to MCI/ADD, be older, and have ≥1 APOE ε4 alleles, compared to AD- and Intermediate statuses (Table 3A).

**Table 3.**
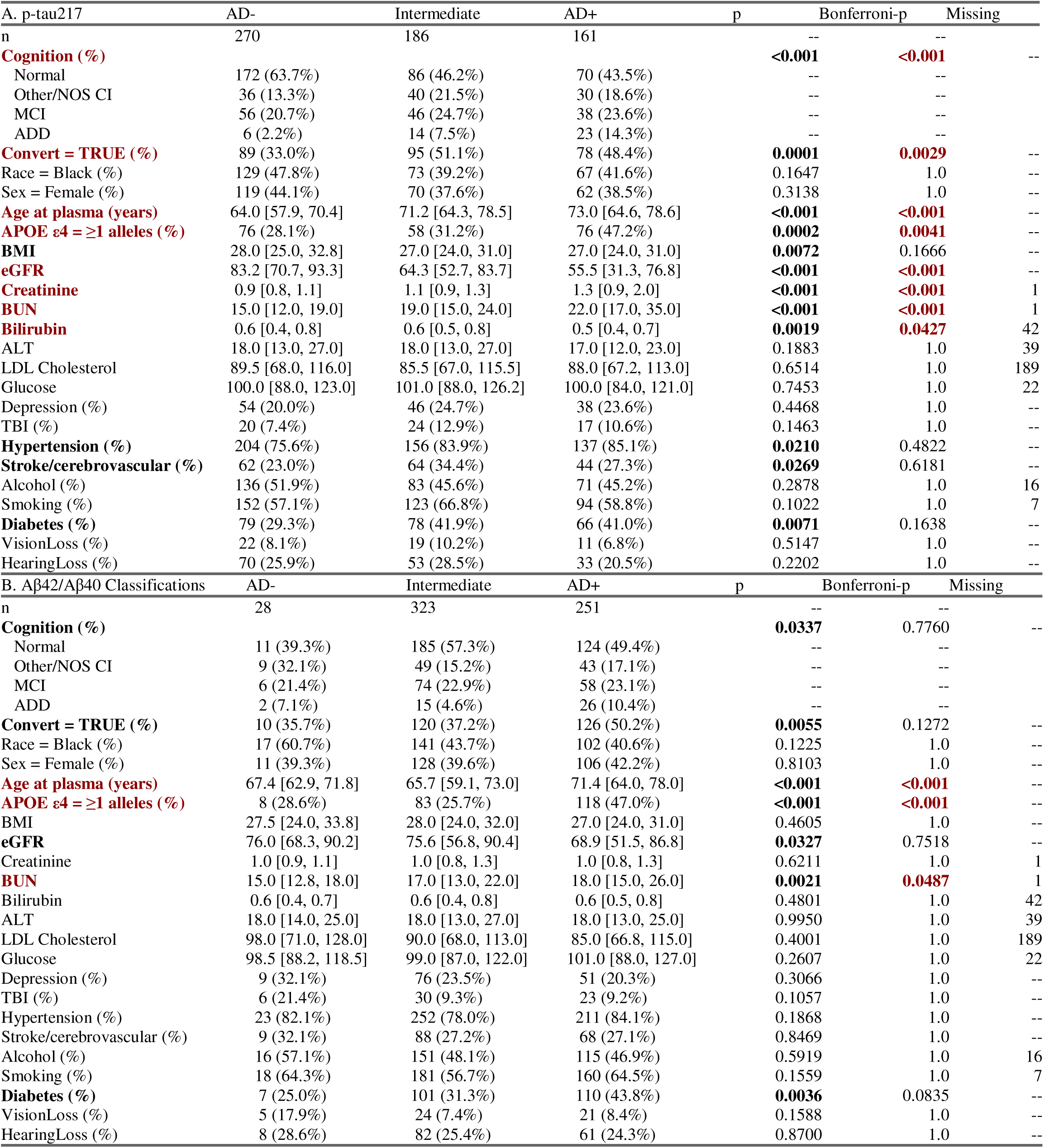
Demographic and morbidity factors that affect plasma p-tau_217_/A. β**_42_ classifications**. Summary statistics across AD negative, Intermediate, and positive classifications. Kruskal-Wallis and Chi-square tested group differences, *p*-values and Bonferroni-*p* are reported: *p*<0.05 are bolded and Bonferroni-*p*<0.05 are in red.

AD+ status for p-tau_217_ was highest for co/multimorbidities (Table 3A). AD+ had the lowest median eGFR, lowest bilirubin, highest creatinine, and highest BUN, and rates of hypertension compared to AD- and Intermediates. AD+ and Intermediates had lower BMI and higher rates of diabetes than AD-. Intermediates had the highest rates of stroke/cerebrovascular disease.

#### 3.3.3 Plasma Aβ_42_/Aβ_40_, Aβ_42_, & Aβ_40_

##### Concentrations

Figure 5A shows plasma p-tau_217_/Aβ_42_ concentrations by cognitive diagnosis and future progression to MCI or ADD. In multivariable models (Figure 3B), lower Aβ_42_/Aβ_40_ was associated with future progression to MCI/ADD (β= −0.23, 95%CI= −0.42 – −0.04, p=0.018), older age (β= −0.12, 95%CI= −0.21 – −0.03, p=0.007), APOE ε4 carriers (β= −0.23, 95%CI= −0.39 – −0.06, p=0.007), higher bilirubin (β= −0.15, 95%CI= −0.23 – −0.07, p<0.001), and a history of diabetes (β= −0.38, 95%CI= −0.55 – −0.20, p<0.001). In univariable models, ADD (β= −0.32, 95%CI= −0.64 – −0.00, p=0.049), eGFR (β=0.10, 95%CI=0.02 – 0.18, p=0.016), and BUN (β= −0.08, 95%CI= −0.17 – −0.00, p=0.038) were significant (Figure 3A).

**Figure 5.**
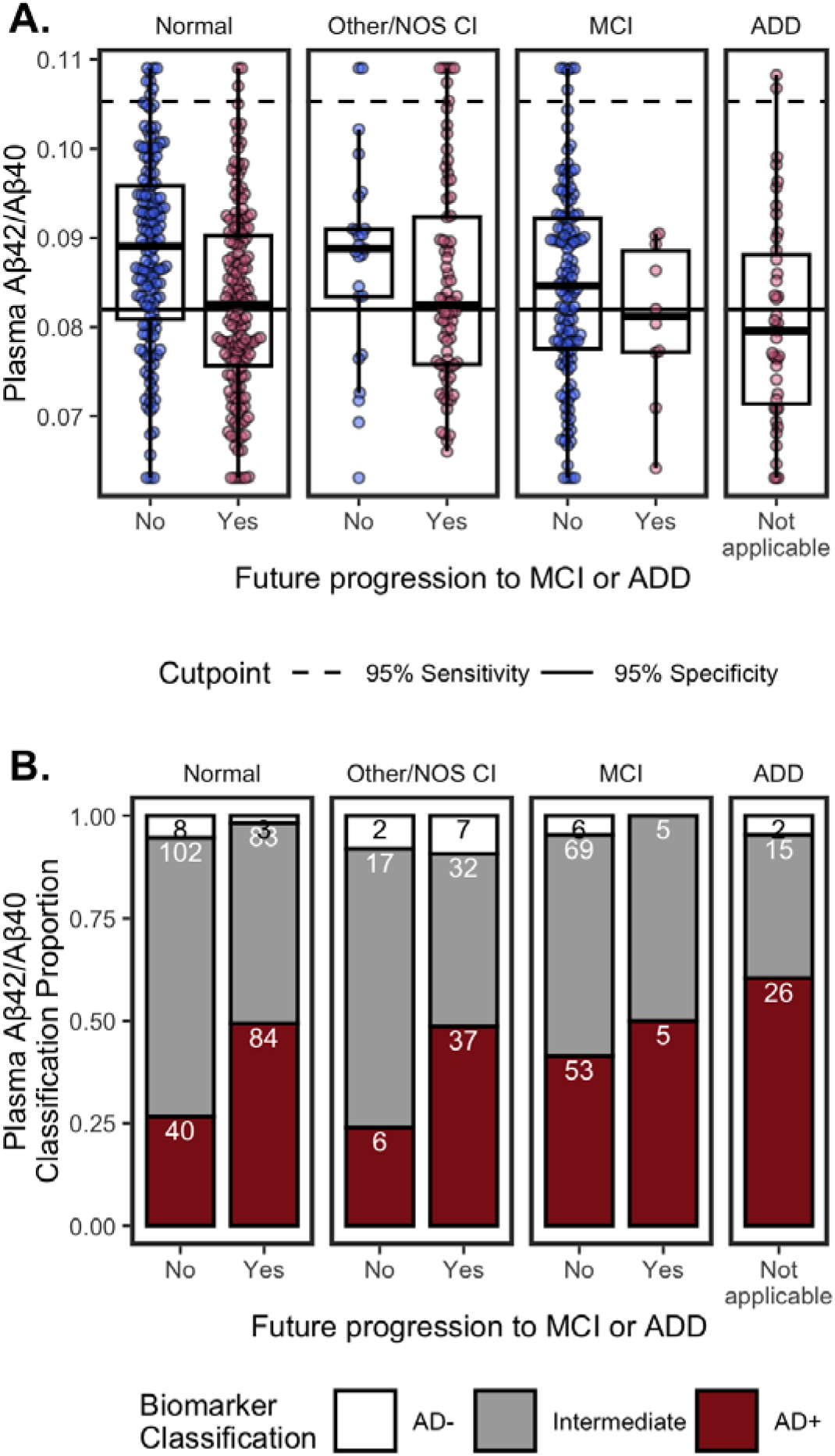
Plasma. Aβ_42_/Aβ_40_ by cognitive status (Normal, Other/NOS CI, MCI, ADD) and future clinical progression. (**A.**) Boxplots show median, interquartile range (IQR), and outliers for each plasma biomarker; high and low plasma levels are Winsorized to 95th percentile for visualization. Color indicates future progression to MCI/ADD (red) or not (blue). Panels show cognitively Normal, Other/NOS CI, MCI, and ADD. Solid horizontal line indicates 0.95 specificity threshold; broken horizonal lines indicate 0.95 sensitivity threshold. (**B.**) Classifiation of Aβ_42_/Aβ_40_thresholds by cognitive status (Normal, Other/NOS CI, MCI, ADD) and future clinical progression. Barplots show proportion of classifications as Amyloid-(white), Intermediate (grey), or Amyloid+ (red). Count for each classification are labeled in bars.

Multivariable models also tested individual Aβ analytes. Lower Aβ_42_ was associated with older age (β= −0.17, 95%CI= −0.24 – −0.09, p<0.001), APOE ε4 carriers (β= −0.18, 95%CI= −0.33 – −0.03, p=0.020), higher BMI (β= −0.18, 95%CI= −0.26 – −0.10, p<0.001), and higher eGFR (β= −0.53, 95%CI= −0.63 – −0.42, p<0.001); no other factors were significant (ALT β= −0.07, 95%CI= −0.14 – 0.00, p=0.065; diabetes β=0.14, 95%CI= −0.02 – 0.30, p=0.092; all other *p*≥0.11). Lower Aβ_40_was associated with older age (β= −0.09, 95%CI= −0.17 – −0.01, p=0.030), higher BMI (β= −0.18, 95%CI= −0.26 – −-0.10, p<0.001), higher eGFR (β= −0.49, 95%CI= −0.60 – −0.38, p<0.001), and lower rates of diabetes (β=0.29, 95%CI=0.13 – 0.46, p=0.001); no other factors were significant (ALT β= −0.08, 95%CI= −0.16 – 0.00, p=0.050; all other *p*≥0.14).

##### Classifications

Using previously established cutpoints^8^, Aβ_42_/Aβ_40_ classifications were 251 (42%) AD+, 28 (5%) AD-, and 323 (54%) Intermediate (Figure 5B). The percentage of Intermediate cases was significantly higher for Aβ_42_/Aβ_40_ (χ2(1)=192.47, 95%CI=0.33 – 0.43, p=9.2e-44) than p-tau_217_/Aβ_42_ (16%).

Consistent with having AD pathology, AD+ classifications were more likely to have ADD, progress to MCI/ADD, be older, and have ≥1 APOE ε4 alleles, compared to AD- and Intermediate statuses (Table 3B).

Examining how medical conditions associated with Aβ_42_/Aβ_40_ classifications (Table 3B), AD+ by had the lowest eGFR, highest BUN, and highest rates of diabetes, compared to AD- and Intermediates, although only BUN survived Bonferroni-correction.

## 4. Discussion

With recent FDA approval for plasma p-tau_217_/Aβ_42_^13^, blood biomarker testing eligibility is certain to expand to heterogeneous and medically complex populations; yet most studies have evaluated blood biomarker performance in the setting of specialized memory centers (but see also^14–16)^. Rigorous implementation and interpretation of blood biomarkers for an AD diagnosis requires continued evaluation in real-world data sets that are representative of populations likely to undergo biomarker testing, including individuals with multiple medical morbidities of varying severity^39–41^. Here, we used EHR data to interrogate how plasma biomarker levels and classification are affected by medical and demographic factors associated with increased AD pathology (AD-related factors) and non-specific dementia risk. Because individual factors may be overlapping or correlated, we tested both univariable and multivariable effects. As expected, all three plasma biomarkers (p-tau_217_/Aβ_42_, p-tau_217_, Aβ_42_/Aβ_40_) were associated with known AD-related factors, including clinical diagnosis of MCI and ADD, age, and APOE ε4. In addition, people who went on to later develop MCI or ADD had higher p-tau_217_/Aβ_42_, higher p-tau_217_, and lower Aβ_42_/Aβ_40_ than those who did not progress. Still, the largest effect size for AD-related factors was observed for p-tau_217_/Aβ_42_. Plasma p-tau_217_/Aβ_42_ also had the fewest number of Intermediate cases, important for providing diagnostic clarity for patients and families^18,42^; our previous work indicates that p-tau_217_/Aβ_42_maintains high diagnostic accuracy even with the small proportion of Intermediates^39^. In sum, our findings demonstrate advantages of the plasma p-tau_217_/Aβ_42_ratio to detect AD pathology in a real-world data set with various medical morbidities, compared to p-tau_217_ or Aβ_42_/Aβ_40_.

Still, p-tau_217_/Aβ_42_ and p-tau_217_ were associated with medical conditions that may confound their interpretation. Foremost, kidney dysfunction – indicated by higher creatinine, BUN and lower eGFR – was consistently associated with higher levels of p-tau_217_/Aβ_42_, p-tau_217_, Aβ_42_ and Aβ_40_. This is corroborated by numerous studies which find correlations between eGFR/creatinine and plasma AD biomarkers independent of AD pathology^23,43,44^. More complicated still, individuals with CKD commonly have other risk factors for AD^45^. Consistent with previous reports, only the Aβ_42_/Aβ_40_ratio was not influenced by kidney dysfunction in multivariable models^43,44^. Still, the effect size of eGFR was somewhat reduced in p-tau_217_/Aβ_42_ compared to p-tau_217_. BMI (obesity), ALT (liver) and histories of diabetes and hypertension were significant univariable factors associated with p-tau_217_ levels, while only diabetes was significant for p-tau_217_/Aβ_42_. Critically, while several medical morbidities (eGFR, ALT, diabetes, hypertension) were associated with AD+ classification by p-tau_217_, morbidities (eGFR, ALT, diabetes) were typically most frequent/severe for Intermediate classification by p-tau_217_/Aβ_42_. While the accuracy of these classifications was not validated in this analysis, it is a notable strength that factors that may confound AD biomarkers are likely to be classified as Intermediate by plasma p-tau_217_/Aβ_42_, which can appropriately prompt further testing^18^. Still, we note that AD+ by plasma p-tau_217_/Aβ_42_ also had more frequent severe morbidities than AD-. Together these results indicate that ratios may help mitigate the confounding effects of medical morbidities on plasma AD biomarkers^5,29,39^.

We found a strong and consistent effect of eGFR on plasma analytes, even in young adults with normal eGFR levels. This may be due to the filtration role of kidneys to clear waste products from the blood stream^37,46^, with better filtration leading to lower analyte concentrations. To account for this effect, we computed eGFR-adjusted plasma p-tau_217_/Aβ_42_ and p-tau_217_. Models showed this adjustment successfully eliminated effects of eGFR, BUN, and diabetes on plasma levels, with no compromise to model estimates of known AD-related factors (effect sizes for ADD and APOE ε4 were modestly/non-significantly increased). Still, it is critical to consider the clinical context for using eGFR-adjusted values, and it is not yet clear if adjusting for eGFR will improve blood biomarker accuracy. Indeed, previous studies find the effect of eGFR/chronic kidney disease on plasma p-tau_217_ and p-tau_181_ is less clinically relevant when AD pathology is high and that adjustment shows negligible model improvement^43,44^. Even so, real-world clinical settings will likely include more persons with severe kidney dysfunction, as well as more various etiologies of cognitive impairment and lower overall AD prevalence^17,18,22^. In this context, kidney dysfunction may result in more false-positive errors when applying biomarker cutoffs. Conversely, our previous study found that false-negative classifications had significantly lower creatinine (*i.e.*, better kidney function) than correctly classified cases^8^. Thus, plasma concentrations may be altered across the full range of kidney function. Still, the value of eGFR-adjusted plasma values to reduce diagnostic errors remains undetermined. Continued efforts to test real-world populations with gold-standard pathology are needed to establish whether eGFR-adjusted plasma values improve classification accuracy and interpretation.

Hearing loss was also associated with lower p-tau_217_/Aβ_42_ and p-tau_217_ in multivariable models after adjusting for age, but not univariable models. Hearing loss is a common chronic condition in aging^47^. Thus, hearing loss had an inverse association with p-tau_217_/Aβ_42_ and p-tau_217_ after adjusting for age, affirming it as either a non-AD dementia risk factor or a confounder to clinical diagnosis^26^.

## Limitations

While the current data generally support the robustness of plasma p-tau217/Aβ42 in a diverse “real-world” dataset, there are several caveats to our findings.

First, this study uses EHR data to investigate the factors of multiple medical conditions on plasma AD biomarkers, and neither formal diagnoses nor neuropsychological testing were available. Likewise, because of this, we are unable to determine diagnostic stability or if there is reversion (*e.g.*, MCI to cognitively normal). Thus, ICD codes should be considered probabilistic of a condition with some error. In addition, continuous or semi-quantitative metrics of disease severity – instead of present or absent ICD codes – may have better power to observe relationships with AD-risk or with plasma biomarker levels. Second, we did not have gold-standard data (*e.g.,*, PET, cerebrospinal fluid) to test accuracy of blood biomarkers in people with co/multimorbidities, or if eGFR-adjusted plasma AD biomarker values improved classification accuracy. Gold-standard data is also needed to confidently disentangle AD-independent effects from factors that increase AD risk. Even so, our findings and others’ suggest that eGFR and creatinine associations with plasma AD analytes are independent of AD pathology^23,44^ and should be accounted for when interpreting biomarker results. Third, eGFR was calculated using creatinine. There is evidence that cystatin C or eGFR calculated using cystatin C are better estimates of kidney dysfunction^48^. Finally, there is a lag between medical screening tests (*e.g.*, eGFR, BMI) and plasma biomarkers that may affect correlations, and incomplete data for LDL cholesterol may have hampered our ability to detect its association with plasma biomarkers.

## Data Availability

Individual-level data from the PMBB are subject to institutional data use agreements and patient privacy regulations, and access is limited to qualified researchers via the PMBB application process.

https://pmbb.med.upenn.edu/investigators.php

## Acknowledgments

We would like to thank the patients and families for contributing to our research. We acknowledge the Penn Medicine BioBank (PMBB) for providing data and thank the patient-participants of Penn Medicine who consented to participate in this research program. We would also like to thank the Penn Medicine BioBank team and Regeneron Genetics Center for providing genetic variant data for analysis. The PMBB is approved under IRB protocol# 813913 and supported by Perelman School of Medicine at University of Pennsylvania, a gift from the Smilow family, and the National Center for Advancing Translational Sciences of the National Institutes of Health under CTSA award number UL1TR001878.

## Consent Statement

Written consent was obtained according to the Declaration of Helsinki and approved by the UPenn Institutional Review Board.

## Data Access

KAQC, CM, AV and Penn Medicine BioBank had full access to all the data in the study and take responsibility for the integrity of the data and the accuracy of the data analysis.

## Author Contributions

KAQC, RB, LMS, and DAW had major contributions to the conception of the research question and overall design of the study. KAQ, RB, CM, AV, CAB, KSO, MS, ND, Penn Medicine BioBank, CTM, EBL, LMS, and DAW were involved in data acquisition and analysis. KAQC performed all statistical analyses and comparisons in this study. KAQC was involved in initial manuscript drafting. KAQC, DAW, and LMS were all involved in the manuscript drafting and editing, and all authors have approved the final draft.

